# Linking private health sector to public COVID-19 response in Kisumu, Kenya: Lessons Learnt

**DOI:** 10.1101/2022.03.29.22271489

**Authors:** M. Omollo, I. A. Odero, H.C. Barsosio, S. Kariuki, F. ter Kuile, S.O. Okello, K. Oyoo, A. K’Oloo, K. Otieno, S. van Duijn, S. N. Onsongo, N. Houben, E. Milimo, R. Aroka, F. Oluoch, T.F. Rinke de Wit

## Abstract

**Background:** COVID-19 is overwhelming health systems universally. Increased capacity to combat the epidemic is important, while continuing regular healthcare services. This paper describes an innovative Public Private Partnership (PPP) against COVID-19 that from the onset of the epidemic was established in Kisumu County, Western Kenya.

**Methods:** An explanatory research design was used. Qualitative in-depth interviews (n=49) were conducted with purposively selected participants including patients, health workers, and policy makers. Thematic analysis was undertaken on interview transcripts and triangulation was performed.

**Results:** The PPP hinged through the provision of central diagnostic COVID-19 services through a parastatal institute (KEMRI). Complementary tasks were divided between Kisumu Department of Health and public and private healthcare providers, supported by an NGO. Facilitators to this PPP included implementation of MoH Guidelines, digitalization of data, strengthening of counseling services and free access to COVID-19 testing services in private facilities. Barriers included, data accessibility, sub optimal financial management.

**Conclusion:** Coordinated PPP can rapidly enhance capacity and quality of COVID-19 epidemic management in African settings. Our PPP model appears scalable, as proven by current developments. Lessons learnt from this initial PPP in Kisumu County will be beneficial to expanding epidemic preparedness to other Counties in Kenya and beyond.

## Introduction

Severe acute respiratory syndrome coronavirus 2 (SARS-CoV-2) has affected the entire world, causing COVID-19, which was declared a pandemic by the World Health Organization (WHO) declared on 11^th^ March 2020 (1). Increased Infection prevention measures (IPC) were recommended, including social distancing, quarantaine, extended COVID-19 testing, distribution of masks to the population and personal protective equipment (PPE) to health professionals and encouraging Water, Sanitation and Hygiene (WASH) Practices, both at community level and at health facilities (2).

As of April 2020, there were about 10,000 confirmed cases of COVID-19 in Africa with at least 500 deaths (3). WHO was putting efforts to mitigate effect of the pandemic on Africa, which was expected to be substantial, given its fragile health systems (4). The first case of COVID-19 in Kenya was confirmed in mid-March 2020 with more cases being reported in Mombasa and Nairobi in April that year (5).

The Ministry of Health (MOH) in Kenya immediately developed an action plan on combating COVID-19 that included restriction of movement, closure of schools, observation of IPC measures, curfews, information campaigns (6). Nevertheless, substantial challenges were encountered, which included limited diagnostic testing capacity, limited availability of Personal Protective Equipment (PPEs), overstretched workforce, weak contact tracing systems and limited coordination of data collection (7).

In order to strengthen the response against the pandemic in Kisumu County and based on previous experiences combating HIV, the Dutch NGO PharmAccess Foundation initiated strengthening a PPP with Kisumu County Department of Health (DoH), Kenya Medical Research Institute/Centre for Global Health Research (KEMRI/CGHR), and key private healthcare facilities through a project named ‘the COVID Diagnostic Project (COVID-Dx)’.

The main aim was to form collaborative and coordinated responses by both private and public sector against the pandemic. Our PPP built capacity at selected private health facilities to complement ongoing public sector efforts to enable more patients in Kisumu County access COVID-19 services. The capacity building was done through training participating health facilities on: COVID-19 clinical screening, coordinated data entry, patient sample collection, safe storage and transportation of samples to the central testing facility (KEMRI). The entire chain of COVID-19 services was supported by digitalization and semi-real time dashboards to keep overview of the entire process.

The current paper provides the results of an extensive feasibility and acceptability study on this unique PPP to provide evidence on the ‘do’s and don’ts of such approaches in times of pandemics. The objective is to describe the experiences and lessons learnt during the implementation of this PPP and probe for its scalability and sustainability.

The three main ‘stakeholders’ in the COVID Diagnostic project were tasked with different roles. The Kisumu County Department of Health owned the project and were taking lead in supportive supervision, setting of guidelines for patient testing eligibility, providing the legal framework. Kenya Medical Research Foundation/Centre for Global Health Research (KEMRI/CGHR) were tasked with conducting operations research together with performing laboratory tests on the COVID-19 study samples. PharmAccess Foundation provided grants for the project, extensive management support, trainings, counselling and contact tracing support, co-creation of digital tools and dahsboards, advocacy and policy makers assistance.

## Methods

### Study Setting and Population

The study was conducted in Kisumu East, Kisumu West and Kisumu Central sub-counties within Kisumu County, Western Kenya. When the project begun, there were seven participating health facilities, of which six remained throughout the entire program and one was dropped due to failure to adhere to some of the Standard Operating Procedures.

COVID-Dx healthcare providers were selected under the existing agreement between the Kisumu County Department of Health and PharmaAccess Foundation based on essential criteria, and non-essential. The essential criteria included: provider should have a license, a COVID-certificate (if applicable), be connected to SafeCare4COVID, reasonable geographic distance within Kisumu County from KEMRI-CGHR laboratories or any future MoH approved testing centre, facilities with ∼100 patients per week, providers (and patients) connected to M-TIBA, average 25 staff, working and serviced fridge and generator for sample storage and willingness and participate in COVID-Dx project. The four additional non-essential but preferred criteria include: high scores on SafeCare, preferably actively using M-TIBA, preferably participating/interested in Medical Credit Fund loan program and participating in PharmAccess MomCare program.

The study population were key informants from policymakers, health workers and patients visiting the participating health facilities.

### Study design and procedures

Grounded Theory qualitative research design was used in the study. This involved conducting In-Depth Interviews (IDI) and Key Informant Interviews (KII) to collect stories of experiences. The interviews were conducted within the environs of articipating facilities. Purposive sampling was used to choose information-rich study participants. A total of 50 study participants were selected for the interviews, and 49 participated (Table 1). The participants were contacted hrough phone calls and emails and written consents were obtained. There were 40 IDIs and 9 KIIs. Thematic approach was used to interpret and triangulate findings.

**Table 1:** Study Participants for IDI and KII

| **Health Facility** | **Patients** | **Health Workers** | **Policy Makers** |
| --- | --- | --- | --- |
| A | 4 | 4 | - |
| B | 7 | 5 | - |
| C | 5 | 4 | - |
| D | 5 | 3 | - |
| E | - | 3 | - |
| F | - | - | 9 |
| Total | 21 | 19 | 9 |

### Data collection and analysis

Trained qualitative interviewers conducted IDIs and KIIs using an interview guide that explored the participants’ opinions on private public collaborations, COVID-Dx services, and COVID-Dx scale up. The interviews were conducted in either Luo, Swahili or English and were audio-recorded. All confidentiality and privacy codes were observed.

Verbatim transcription was done with quality check performed on all scripts. A thematic framework was developed in line with the study objectives alongside the data on the transcripts. The thematic framework was used to develop a codebook in Nvivo 11. Coding was done on the transcripts with inter-coder reliability run for quality check. Text matches, coding queries and coding matrixes were run to interpret the coded data further. Findings were reported using graphs and charts.

### Ethical Approval and Consent to Participate

The study received ethical clearances from two bodies: Kenya Medical Research Institute: Scientific and Ethical Review Unit (SERU) KEMRI/SERU/CGHR/05/05/4038: and Jaramogi Oginga Odinga Teaching and Referral Hospital (JOOTRH) Ethical Review Board (ERB) IERC/2030/2020. Additional approval was received from NACOSTI with license number NACOSTI/P/20/5616. Written consent was obtained from all participants before the process of data collection.

## Results

Data was categorized thematically according to each set of respondents i.e Patients, Health Workers and Policy makers as shown in Table 1.

The respondents had various demographic characteristics comprising of gender, age, education, and cadre characteristics. In terms of gender, 23 were male with 26 female with 87% of both respondents having completed at least diploma education. In terms of professional cadre, 40 participants were healthcare providers ranging from Nursing officers, clinical officers, laboratory technologist, health facilities administrators: with 9 participants being the staffs from Kisumu County and Sub Counties Department of Health

### Policy makers’ experiences

Public facilities that were designated to handle COVID-19 in Kisumu County were initially few. Support by the private facilities enabled filling in gaps and thus help mitigate the sometimes overwhelming situation in public facilities. The public sector initially took lead with surveillance activities, follow-up and contact tracing of COVID-19 cases. The opportunities that came with private facilities accelerated these services. Public facilities were initially better positioned in terms of trainings and medical supplies. However, they experienced barriers with services, including overcrowding and thus long waiting times for patients:

> “*The issue of overcrowding, the overcrowding aspect I think when you came in you were of great help to us really. Because initially the testing facilities were only…I remember they were only two. We were really overwhelmed and I remember there were only two private facilities doing this. For the public facilities we were only having two facilities that is District and JOOTRH, you can imagine it was covering the city life ‘that’s why partly the Kisumu west, partly Kisumu East and partly Kisumu central. So we were really overwhelmed when it comes to testing*.*” KII 3 Female*

The private sector was facilitated to be fully aligned with public interventions, including usage of existing protocols that governed rollout around results management, information delivery, coordination and management of health systems in general. There was a staff gap filled by involving the private facilities in scheduled activities organized by the public facilities such as trainings, outreach and couseling. There was alignment of overall COVID-19 testing capacity, definition of the criteria qualifying a patient for such a test, organizing logistics of dentralized testing through provision of trained (motorbike) transport. Moreover, public and private data collection became much better coordinated, facilitated by customized software and hardware: tablets were provided to all stakeholders in the chain of sample and data flows to secure flawless tracking and tracing. Together the PPP identified the need of a disease response team, with representatives from both public and private facilities. All these items were addressed through dedicated trainings of PPP stakeholders, as facilitated by the NGO.

> *“One thing I realized is that when we began training, I think we trained the public facilities first. Little did I know that the private facilities were actually on board until we began training the private facilities when I realized, wow so you mean we’ve not been able to train the private facilities all this time? So, I think there’s some kind of de-link that needs to be worked on such that in case we have such kind of scenarios next time then everybody needs to be brought on board almost immediately”. KII 1 Male*

Capacity building of health workers and facilities and increased testing in the private facilities enabled health seekers to go for the COVID-19 services at these facilities thus reducing the workload on the few public facilities that were offering COVID-19 services.

> *“It is good, I mean it’s good. Like now in Kisumu they’re helping, like we have cases right now in (Name withheld), we have cases right now in (Name withheld). So, they’re actually helping; if we left this for the public facilities only: (Name Withheld) and (Name Withheld), I don’t think we’ll be able to cope with the challenge that is there. “KII 1*

Scaling up the COVID-Dx services through understanding their compatibility with the MOH Guidelines on COVID-19 management was key. COVID-Dx was co-created in full collaboration with the Kisumu DoH. COVID-Dx contributed to training of health care workers, provision of complementary PPE, sample collection and transportation process, data management and creation of digital dashboards for semi-real time monitoring and evaluation. The strengthening was in terms of bringing on board and capacitating the private health facilities. General funders of healthcare like private insurance companies, public National Hospital Insurance Fund (NHIF), bilateral donors did not cover COVID-19 treatment services. Funding for COVID-19 in the public facilities was facilitated by the MOH through provision of consumables and monthly allowances.

> *“Well, I was impressed because the COVID-Dx had a design that was easily entrenched towards; I saw it as something that was strengthening our existing system. So, it got in very smoothly and you could not know that it is a private partnership or tri-partnership the way it was. Because it works as a unified agency. You could think that it is run by the government, but you see; it is three*.*” KII 6 Male*

> *“I think COVID-Dx has always been operating under the MOH guidelines. So they conform, they don’t have any other guideline but they conform to the MOH guideline in conducting the activities. There is nothing they do that is different because they collect samples as required by the MOH. They enroll participants in line with what the MOH stipulates. The testing is done at the central laboratory at KEMRI where the MOH does theirs, the results are disseminated to the MOH so as much as the study gets the results they go through the MOH*..*” KII 8 Male*

Scaling up and integration of COVID-Dx approach in the county’s health system was key to fill in the existing gaps including curbing challenges on adaptation of the use of technology which comes with increased transparency, accountability, capacity building, networking and increased access to testing services. Facilitators to scaling up and integrating COVID Diagnostic Project approach included clear guidelines and policies for all the parties involved in the scale up. Barriers mentioned included limited funding, wider coverage, understaffing of private facilities and mismanagement of funds in public facilities

> *“Well, I think it should be …*.. *because there have been these counties that have been hard hit and I think integrating this, because that is what I’ve also learnt over the years; that an integrated approach towards management of diseases is the best approach. “KII 9 Male*

PPP needs coordination. In order to achieve this, all partners should be brought on board to participate in putting together the resources and manpower in bridging in the gap between private and public facilities. A third-party facilitator (such as PharmAccess) can truly catalize this.

> *“So I think my recommendation would be private-public partnership should be on the table from day one. They should not come later. The private sector must be there when a pandemic is announced, apart from having all government state organs on board. All stakeholders must be there and they must be treated as equal partners. No matter how small a health facility is, it must be treated as a partner. KII 5 Male*

### Health workers’ experiences

The respondents experienced new opportunities through the PPP including capacity building of private facilities in terms of training and availing commodities, referrals, and surveillance for COVID-19 service provision. The few risks that were experienced, were during project’s onset where the facilities feared being closed down in case they had COVID-19 positive clients or staff. These fears disappeared rapidly.

> *“Our collaborations with the public health facility with the working in conjunction with the public health facilities have assisted us getting essential supplies for testing, like viral transport medium (VTM) and swabs and gowns and everything. So collaborating with the public health facility has assisted us to achieve whatever we wanted to with this COVID 19”*.*” IDI 12 Facility C, H/W*

On the experience with COVID-Dx services, the trainings offered on IPC, sample collection and transportation were considered most efficient and educative. Still there was need to have frequent refresher trainings in addition to certifications.

PPEs were availed timely, used and discarded with only recycling being done to the goggles, face shields and the plastic perforated footwear after disinfecting. There were instances where health workers shared concerns about poor quality of PPEs, especially when it came to the doffing process that poses an avenue for contamination.

“*What I would like to share is that I did not know the quality of PPE. So that one maybe during the training would be sure this is the right quality, this one is approved by KEBS. So maybe quality depends with those who supply them to the sites for use. So next time they improve on quality because… and again if they can make it better so that we don’t sweat. There is serious sweating in that PPE*.” IDI 1 Facility A, H/W

The health workers were enlightened with the opportunity for personal diagnostic screening that the project offered to them. This was done on a 2-months-basis, but in cases where the health workers were exposed or felt symptoms they had access to immediate testing. The process of sample collection after training went well, especially with oral-pharyngeal samples. Nevertheless, health workers experienced the collection of samples as stressful and asked for hazard allowances. These were not provided, but care was taken to maximize observation of IPC measures.

> *“The screening is okay because if I can say, like if we as the providers anytime we get a positive case we do for the tests. After maybe three days we go, no five days we go for the test. But routinely if we ‘don’t have any positive case, we like do it twice in a month”*.*” IDI 19 Facility B, H/W*

After collection, samples were packed and transported to the reference laboratory immediately. There were few cases that the sample had to stay for longer hours at the facility in the fridge. The majority of the results were reported back to the facility between 24-48 hours. The facility then communicated the results to the patients. In few instances, there could be delays in results communications. Initially, patient management was done by the Kisumu County DoH. However, as COVID-19 infection went up, contact tracing and patient management became a challenge to the Kisumu County DoH and was taken over by the participating health facilities.

> *“We have not had a challenge with sample storage and transportation. Apart from the delays we are good”*.*” IDI 30 Facility E, H/W*

> *“So in terms of diagnostics’, we’ve had good support from the public sector, but in terms of referral, there were quite a lot of challenges. Moreover, we have this home-based care which permits for those who are stable. We have to involve the public officers to access these clients’ homes who are to be released for home-based care. It is not every time the public officers are readily available to do that. So at the end of the day home based care and referral to public facilities for COVID patients has been a big challenge*.*” IDI 31 Facility E, H/*

### Patients’ Experiences

The experiences with Dx services were positive. The respondents noted that when they visited the COVID-Dx facilities for sample collection, IPC measures were being observed from the facility entry, triage desk, ‘clinician’s room and sample collection point with health workers in their PPEs.

> *“The procedure was okay I was explained to how it would be. I was told it would be somehow uncomfortable. Of which indeed it was somehow uncomfortable but something you can bear. Basically there was no inconveniences”*.*” IDI 40 Facility D, P/T*

The sample collection process was experienced as uncomfortable especially the naso-pharyngeal method, and most of the respondents were experiencing teary eyes with few sneezing. Respondents shared their satisfaction in the way the results were conveyed, with few respondents mentioning some of the inconvinie experiences they had with results delivery considering the timing and the observation of the confidentiality aspects.

> *“Since I was told 24 hours, I expected the following day I will get my results because I was told I will be given a phone call. So I waited for the phone call but I did not get it. So I’m the one who decided to start making the phone calls. I called and called till I went there, they told me to go to (Name withheld) for my results. When I went there they told me they can’t give them to me. So they gave me the county officer’s number who had the results. So when I called him, he told me he is in the field and I should not go to the county office and that I should go wait for him at the district hospital*.*” IDI 16 Facility C, P/T*

Patient management was mostly by home-based isolation for the asymptomatic patients. Few respondents were not in favor of quarantine. Some respondents shared concerns about non-observance of IPC measures, lack of provision of health education on COVID-19 as they were waiting to take the test, and delays in conveying test results.

> *“They have sanitizers, water and soap at the ‘clinician’s rooms, which is good. But what can be checked on that room is: sometimes people who are stubborn and ‘don’t want to put on their masks enter into this room and ‘he’s attended to. Sometimes ‘‘‘sick, he’s sneezed and coughed; you know the clinician has the mask on, but you know they said that COVID stays airborne. So, when he leaves this room, the other person ‘that’s coming into this room can be infected”*.*” IDI 14 Facility C, P/T*

## Discussion

This study describes experiences and lessons learnt from a pioneering PPP to combat COVID-19 in Kisumu County, Kenya. Information was collected through 49 In-Depth amd Key Informant Interviews held with the Patients, Health Workers, and Policy Makers. Previous studies have shown that PPPs can have positive impact in healthcare accessibility with major regards to treatment and prevention services (8).

The experiences of all respondents with this PPP (patients, providers, policy makers) were majorly positive. COVID-Dx facilities collaboratively providing COVID-19 services reduced overwhelming workloads at the public facilities and increased opportunities for clients to be tested. The testing capacity increased, as well as trainings, provision of commodities and better integrated surveillance activities. The PPP alignment worked well as the protocol that governed the partnership was in line with the existing MOH guidelines on COVID-19 service delivery. Not all private facilities could always participate in trainings, due to workload, staff turnover and sometimes limited availability of public sector trainers. During the onset of the partnership, some of the private facilities feared being closed down in case they have COVID-19 positive clients.

Sharing of knowledge on COVID-19 was done by regularly bringing together the participating facilities to share their experiences and by organizing trainings and refresher courses. This is along reported experiences that indicate that sharing knowledge and interim research results among collaborators during management of a pandemic provides an important learning forum (9). Training of healthcare workers on how to combat COVID-19 is one of the rapid responses that can be done collaboratively (10).

Experiences with COVID-Dx services that were noted as positive included: training of health workers, timely provision of PPEs, consumables and commodities, better sample collection and transportation with high observation of IPC measures, coordinated and standardized data collection, health workers’ regular COVID-19 screening, timely results management and patient management. There were mixed feelings about the quality of some commodities like PPEs which posed avenues of contamination because of their design. In addition there were complaints about the turnaround time when delivery of test results went beyond the promised 48 hours. The sample collection process was noted to be uncomfortable. Patient management that involved quarantine was not perceived positively. None of the insurances, neither private nor public covered COVID-19 treatment services.

The starting point for COVID-19 PPP was also inspired by the Africa Centre for Disease Control and Prevention (Africa-CDC) and the WHO, both of which advocated for partnerships to accelerate COVID-19 tracing, testing and outcomes in Africa (11). The well-known statement by WHO Director, Dr Tedros: ‘test test test’ was taken as the hinge for welding our PPP COVID-19 response in Kisumu (12). Particularly at the onset of an epidemic, collecting data on its geographic spread, target population is crucial and therefore diagnostic testing has priority. This was applied in the current PPP in Kisumu by linking both public and private healthcare facilities to a centralized parastatal testing facility (KEMRI). Thus, the workload on the public sector was alleviated, while at the same time the private sector was educated about public sector clinical guidelines, testing eligibility criteria, result reporting and patient tracking and tracing mechanisms.

A key lesson was the crucial role of a ‘third party’ entity (in this case an NGO, PharmAccess Foundation) to facilitate collaboration between public and private sector. This approach alleviated the extra efforts that otherwise would have to be delivered by already overwhelmed public and private healthcare staff. With limited funding the ‘third party’ approach was able to quickly accelerate COVID-19 responsiveness in Kisumu. This was particularly achieved through a flexible ‘can do it’ approach, where gaps identified either in public or private healthcare delivery were temporarily filled in by the ‘third party’ and subsequently training was provided to help either public or private sector, or both to fill the gaps.

The success of this PPP in Kisumu has not gone unnoticed and the epidemic preparedness capacity built proved to be of great importance in next developments. First of all, this PPP, despite all its efforts, led to the experience of general lack of SARS-CoV-2 testing capacity through PCR. Therefore, an intervention was established introducing for the very first time rapid diagnostic COVID-19 tests and validating their performance in Kisumu field-setting (13). This has opened the possibility for private facilities to complementarily procure rapid tests in relatively smaller quantities at affordable prices and thus be less dependent of government supplies and in general increase the COVID-19 testing capacity for Kisumu citizens. Subsequently, at the (financial) closure of this project, in April 2021, a sudden outbreak of COVID-19 Delta variant was experienced in Kisumu (14). This started in a sugar factory with Indian workers and spread quickly to the city of Kisumu. Immediate action was required to identify the infected and try to ring-fence the epidemic. Based on the trust and collaboration that was built through COVID-Dx, the DoH of Kisumu requested immediate implementation and scaling. With emergency funding this was realized and within weeks, the existing COVID-Dx PPP infrastructure was expanded from 9 to 32 facilities, including trainings, customized data entry systems and dashboards to keep overview. This project will be described in a separate paper. Lastly and very recently, the Lake Region Economic Block, a consortium of 14 Counties in West Kenya has approached COVID-Dx Kisumu to copy its model into the entire LREB area, which is serving one third of the entire Kenyan population. This scaling is currently ongoing, to build a Western Kenya digital epidemic preparedness system, all based on the original COVID-Dx PPP accomplishments.

## Data Availability

All data produced in the present work are contained in the manuscript

## Strengths and Limitations

The strength of the study was its operational nature, flexibility of interventions and the establishment of mutual trust by building compatibility between otherwise relatively isolated private sector and the existing public health system, including the MoH and its guidance on managing the COVID-19 pandemic. The qualitative data collection method of the study enabled interviews with different stakeholders resulting to comprehensive overview of their experiences. Limitations were experienced due to the initial need to position this PPP as a ‘research project’, implying all formal procedures for such an endeavor. Later during implementation this requirement was adjusted.

## Conclusion and recommendations

The experiences and lesson learnt from an innovative PPPP in Kisumu, Kenya, combating COVID-19 pandemic proved vital. The digital infrastructure built could be expanded quickly when the COVID-19 epidemic increased. Such epidemic preparedness will prove its use and efifiency for future outbreaks of COVID-19 or any other epidemic in Western Kenya. It is recommended to involve third party entities to amalgamate and facilitate public-private collaboration, particularly during emergencies, like epidemic outbreaks. In addition, buffer stocks should be established of essential medical commodities and supplies at strategic and safe locations in the country. Moreover, it is recommended to have expedited (legal) procedures ready to support immediate roll-out of epidemic interventions. And finally, there should always be evalution and continued learning around epidemic interventions by involving operational researchers and institutes, analyzing data and making lessons learned available to policy makers.

## Acknowledgements

University of Amsterdam, Netherlands

PharmAccess Foundation, Kisumu

Kenya Medical Research Institute/Centre for Global Health Research

Kisumu County Department of Health

Healthcare providers, patients

## Appendix

**Figure 1:**
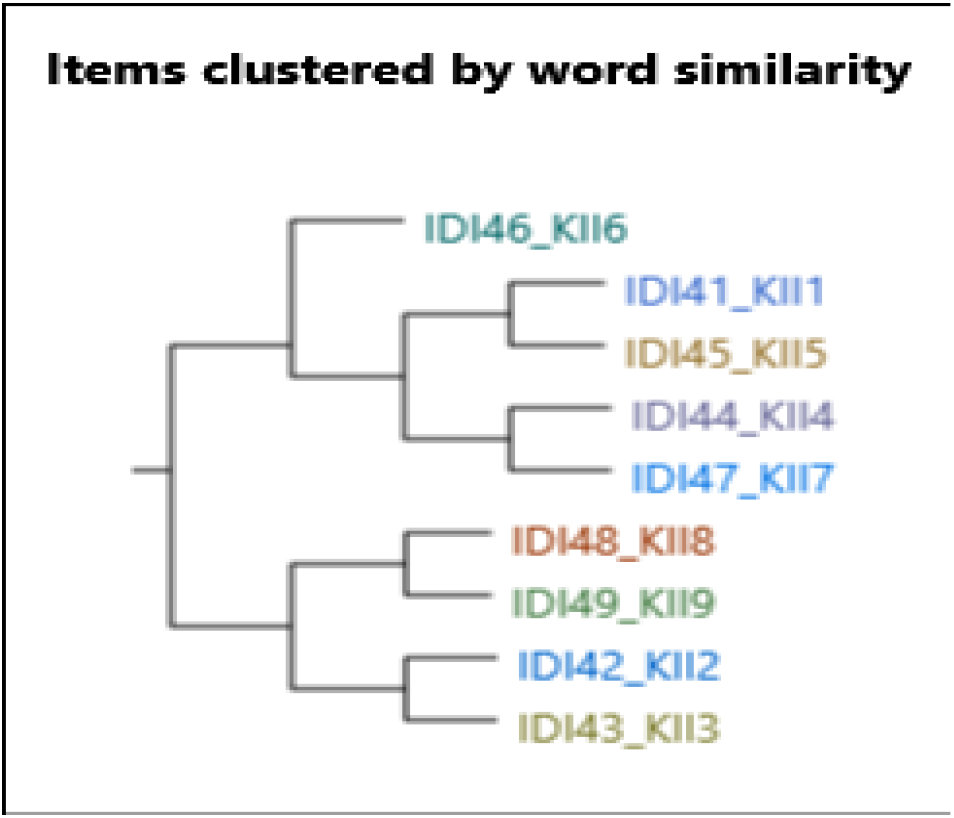
A cluster showing similarity by word contents among the respondents of the Key Informant Interview Category. The categorization indicate the similarity in terms of the contents, like for example respondent 46 had unique content thus there is no any other responses that is muching with. Respondent 44 and 47 had similarity in their contents.

**Figure 2:**
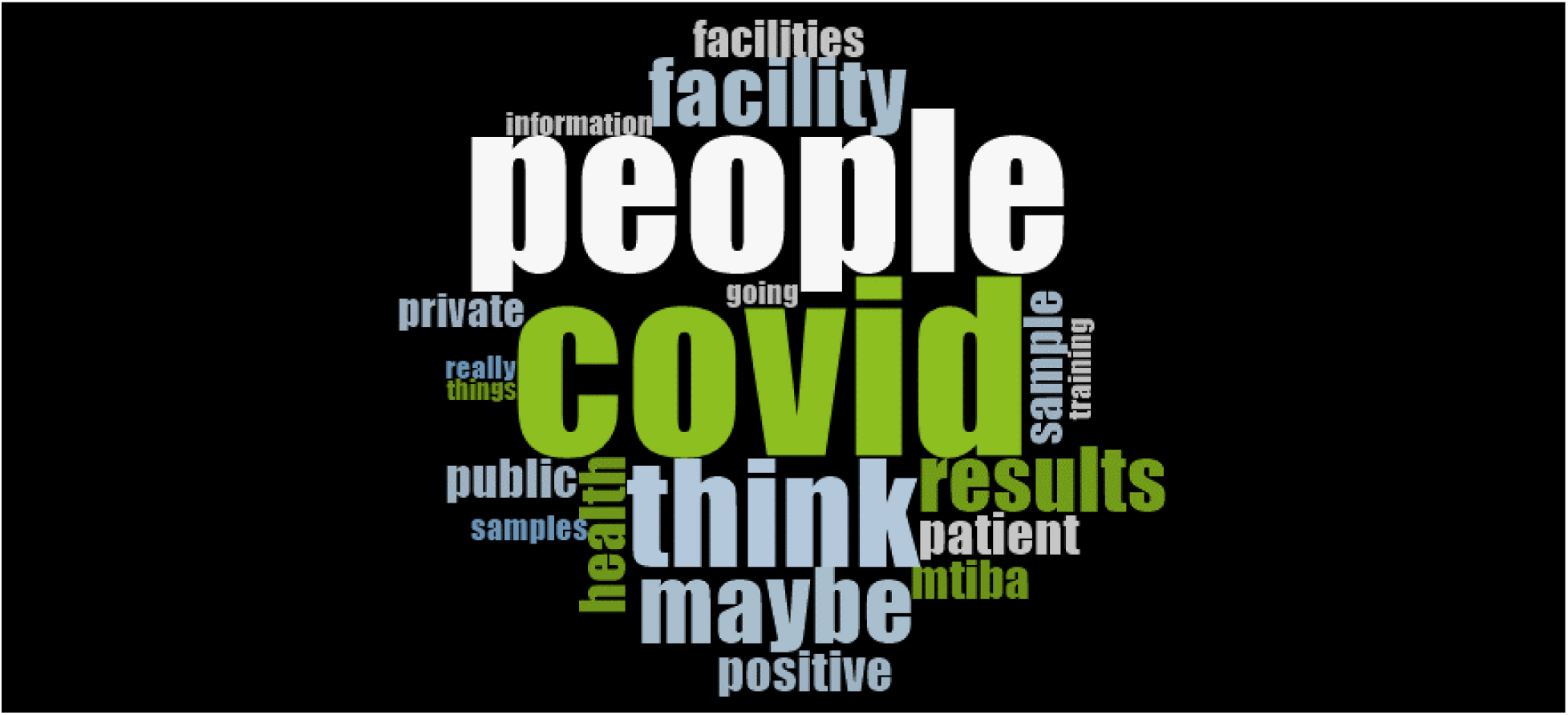
A Word cloud on the most frequently used word by the respondents. This shows the commonly used words and phrases that were mentioned during the interviews.

**Figure 3:**
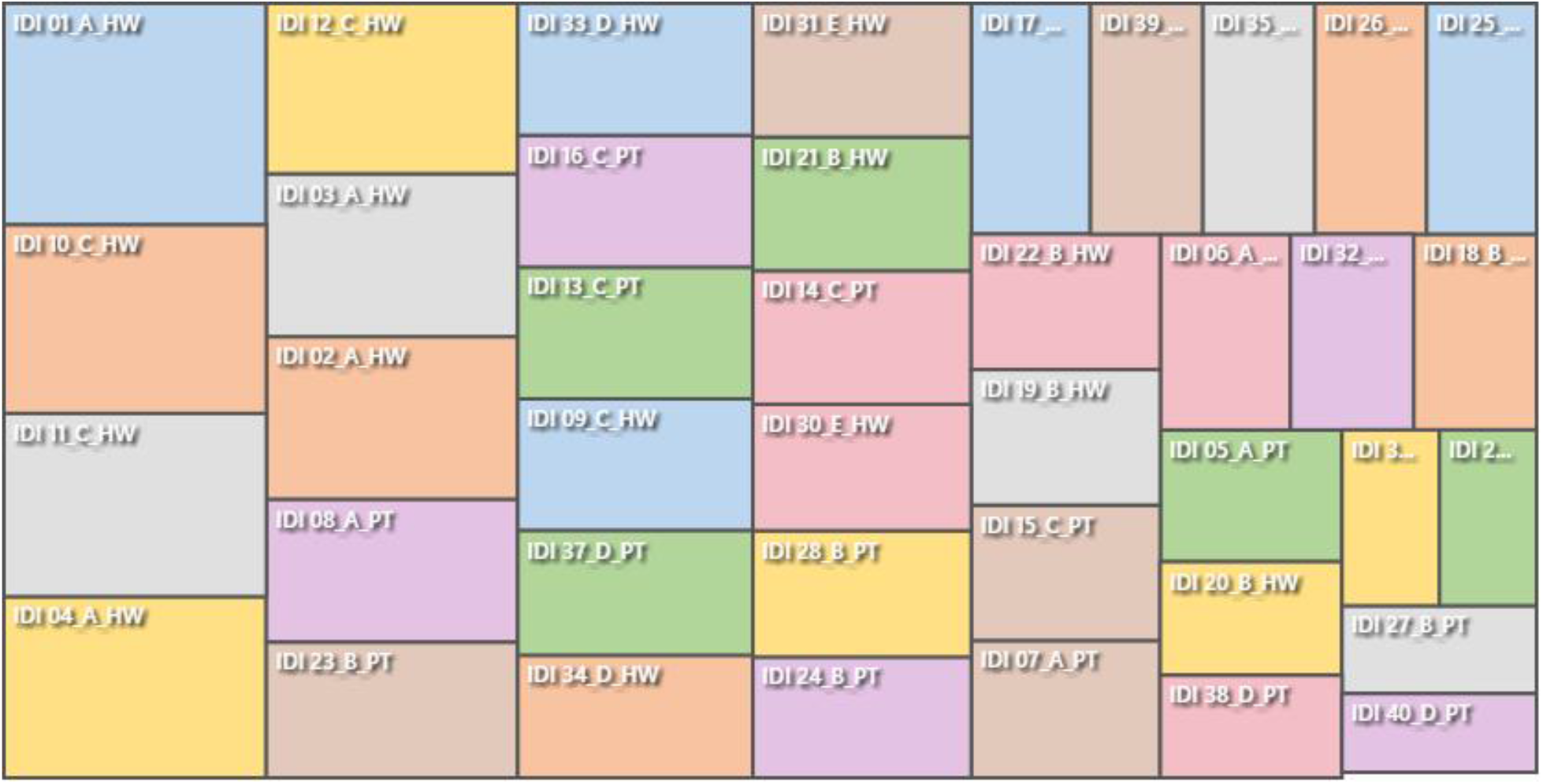
Chart showing respondents experiences with COVID-Dx services. The larger the boxes the higher the level of content shared. For Example, IDI 01 shared a lot about experiences with COVID-Dx services as compared to IDI 34

